# Modelling Negative Symptom Domains Neurobiology: Protocol for an Observational Transdiagnostic, Translational Study

**DOI:** 10.64898/2026.01.24.26344630

**Authors:** Cătălin Dan Oprea, Ilinca Untu, Diana-Sabina Vieru, Camelia Dascălu, Helene Speyer, Cristina Dobre, Ohad Green, Romeo Petru Dobrin, Michael Davidson, Jonathan Rabinowitz, Bogdan Ionel Tamba

## Abstract

**Background:** Negative symptoms (NS) such as anhedonia, avolition, asociality, blunted affect and alogia are associated with poor functional outcomes in psychiatric and neurological disorders and are an unmet treatment need.

**Objective:** This protocol describes the design of an observational, single-center study aimed at characterizing negative symptoms across a transdiagnostic sample of individuals with mental disorders and related conditions, with a particular focus on avolition, its biological correlates, and associated neurocognitive and electrophysiological profiles.

**Methods:** An observational study (involving no invasive procedures or drug administration other than a routine blood draw) has been designed to examine negative symptoms across psychiatric disorders. A total of 300 participants with a primary diagnosis of schizophrenia, bipolar disorder, unipolar depressive disorder, autism, or dementia will be recruited, with a target of at least 50 individuals in each diagnostic group. Consenting inpatients and outpatients will complete a battery of non-invasive behavioral and cognitive assessments, undergo electroencephalogram (EEG) recording, and provide blood samples for the assessment of polygenic risk scores.

**Results:** The initial version of the study protocol was developed in February 2024. The finalized protocol was completed on August 5, 2024, and subsequently updated on January 11, 2025, to incorporate minor methodological clarifications. Participant recruitment and data collection commenced on July 1, 2024, and are ongoing at the time of manuscript submission. Data quality control and preliminary analyses are performed concurrently with data collection, while final statistical analyses and dissemination of results are planned following completion of the recruitment phase.

**Conclusions:** This study will provide critical insights into the characterization and underlying mechanisms of negative symptoms across psychiatric disorders. By focusing on avolition, reward processing, and their interaction with neurocognitive and social cognitive deficits, it will help identify potential biological and electrophysiological markers of negative symptoms. The findings may guide the development of more precise assessment tools and inform novel therapeutic strategies, with broad translational impact for improving outcomes in individuals with serious mental illness and related conditions.

## Introduction

Mental disorders represent a major global health burden, being among top ten leading causes of disability [1]. Depression and anxiety are the most prevalent conditions, while psychotic disorders, such as schizophrenia, affect approximately 0.33% to 0.75% of individuals worldwide [2-4].

Negative symptoms represent a core dimension of psychopathology across the spectrum of serious mental illnesses, including schizophrenia, bipolar disorder, major depressive disorder, and neurocognitive disorders. Unlike positive symptoms, which reflect an excess or distortion of normal function, negative symptoms involve a reduction or loss of fundamental psychological processes [5]. Within severe mental illnesses (SMI), such as schizophrenia, bipolar disorder, and unipolar depressive disorder, as well as in neurocognitive disorders including dementia, negative symptoms (NS) remain a significant unmet treatment need [5].

Negative symptoms refer to a reduction or loss of normal psychological functions and are classically grouped into two domains: (1) Experiential domain, comprising anhedonia (reduced ability to experience pleasure), avolition (marked reduction in motivation and a diminished ability to initiate, sustain, and complete goal-directed activities), and asociality (reduced interest in social interactions and diminished desire to form or maintain relationships); and (2) Expressive domain, including blunted affect (marked reduction in the expression of emotions, observed as limited facial expressions, reduced eye contact, monotone speech, and decreased use of gestures) and alogia (poverty of speech and reduced verbal output). Importantly, NS are strongly associated with poor functional outcomes, impaired quality of life, and limited response to current therapeutic approaches [5].

In the neo-Kraepelinian era, psychiatric nosology has largely conceptualized mental disorders as natural kinds, understood as discrete disease entities whose observable symptoms are taken to reflect underlying, latent pathological processes. This framework has shaped diagnostic systems by prioritizing reliability, categorical classification, and the assumption that symptoms cohere because they are manifestations of a single, unifying disorder [6]. However, accumulating empirical evidence has increasingly challenged this view. Research has demonstrated substantial symptom overlaps across diagnostic categories, high rates of comorbidity, and heterogeneity within diagnoses, calling into question the notion that traditional categories map neatly onto distinct biological or psychological mechanisms.

In response to the view that mental disorders are discrete entities, more recent approaches have shifted focus from distinct disorders to symptom domains that cut across diagnostic boundaries. Within this transdiagnostic perspective, symptoms such as anhedonia, anxiety, or cognitive dysfunction are investigated as phenomena in their own right, potentially sharing mechanisms across multiple conditions. Building on this shift, network-based models represent a further conceptual departure by rejecting the assumption of a latent disease entity altogether. Instead, they conceptualize psychopathology as emerging from direct, dynamic interactions among symptoms themselves, such that symptoms mutually reinforce and sustain one another over time. From this viewpoint, mental disorders are not the causes of symptoms but rather the emergent patterns arising from complex symptom networks, with important implications for diagnosis, aetiology, and intervention [7].

Recent advances in behavioural, cognitive and pathophysiological research highlight the need for a more precise characterization of NS to enable the development of innovative treatments. Evidence suggests that, at least in part, NS may share common biological substrates and transdiagnostic manifestations. Repeated analyses of neural networks involved in emotional and cognitive processing indicate that avolition is a central component of NS, playing a role in the generation and maintenance of other symptoms. Moreover, improvement in avolition is often accompanied by amelioration of the broader spectrum of negative symptoms [8].

Network theory offers a useful framework for advancing the understanding of negative symptoms as transdiagnostic phenomena because it shifts the focus from diagnostic categories to the dynamic relations among symptoms. Network models allow these symptoms to be examined as interacting components of a system rather than as passive indicators of an underlying disorder [9]. By modeling symptoms as nodes and their associations as edges, network analysis can test whether negative symptoms form stable, coherent clusters across diagnoses or whether their organization, centrality and connectivity differ between conditions [7]. This approach makes it possible to identify symptoms that act as central hubs or bridges linking motivational, affective, cognitive and functional domains, thereby clarifying which negative symptoms may play a driving versus downstream role in different diagnostic groups [9].

In line with the overarching aim of characterising negative symptoms across diagnostic categories, this study further seeks to examine the relative importance of individual negative symptoms within symptom networks by assessing measures of node centrality. Drawing on network-theoretical approaches, centrality indices such as strength, closeness, and betweenness are used to identify symptoms that are most influential within the network of negative symptoms. Symptoms with high strength may exert broad influence through strong direct associations with other symptoms, whereas symptoms with high closeness or betweenness may play a key role in maintaining or propagating symptom dynamics across the network. By comparing centrality profiles across diagnostic categories, the study aims to determine whether particular negative symptoms consistently occupy central positions across disorders or whether centrality patterns differ by diagnosis. This approach directly supports the study’s goal of disentangling shared versus disorder-specific mechanisms underlying negative symptoms and may help identify key targets for transdiagnostic intervention.

### Objectives

The overall aim of this project is to characterize persistent negative symptoms in a transdiagnostic sample of individuals with severe mental illness by investigating the mechanisms underlying different domains of these symptoms. Specifically, the main objectives of this study are: (1) Characterize negative symptoms in a transdiagnostic sample by modelling their interrelations using network analysis, and compare network structure and symptom centrality across diagnostic categories to distinguish shared versus disorder-specific mechanisms; (2) Examine whether *avolition occupies a central* position within negative symptom networks, supporting its proposed role in generating and maintaining other negative symptom domains; (3) Investigate associations between symptom centrality, particularly of avolition, and *biological markers*, including genetic and other biological measures; (4) Examine alterations *in reward anticipation and reward experience* in relation to negative symptom profiles and motivational deficits, and their interactions with neurocognitive and social cognitive dysfunctions; (5) Link network-derived symptom profiles to *electrophysiological measures*, comparing patterns across symptom profiles, diagnostic categories, and treatment response.

Participants will be recruited according to ICD-10 criteria from the following diagnostic categories: schizophrenia (all forms), schizoaffective disorder (all forms), persistent delusional disorder (including the schizophrenia spectrum), major depressive disorder, bipolar affective disorder, autism spectrum disorder (diagnosed during childhood/adolescence), and various forms of dementia. An effort will be made to include a similar number of patients in each diagnostic group; however, depending on availability, group sizes may vary. The estimated cohort for this study is 300 patients. To achieve its objectives, the study will employ a multimodal assessment protocol including psychometric and clinical scales, neurophysiological recordings, and biological sampling.

## Methods

### Study design and setting

This is an observational, single-group, exploratory study. All study visits will be conducted at the Socola Institute of Psychiatry, an academic hospital affiliated with Grigore T. Popa University of Medicine and Pharmacy Iaşi, Romania. Data collection will take place exclusively at this single site. The trial will recruit an estimated 300 participants across multiple diagnostic categories of severe mental illness and related disorders. Participants will be recruited from both inpatient and outpatient treatment. Personal data will be anonymized to ensure confidentiality. An effort will be made to include a similar number of patients in each diagnostic group, aiming for at least 50 subjects per group, with possible increases up to 80 depending on availability. The study framework is exploratory, aiming to characterize persistent negative symptoms, identify potential biomarkers, and investigate underlying mechanisms across diagnostic groups.

### Eligibility criteria

Participants are adults aged 18–65 years who present with a primary ICD-10 diagnosis of schizophrenia, bipolar disorder, unipolar depressive disorder, autism, or dementia with a minimum score of 20 on the negative symptom subscale of the Positive and Negative Syndrome Scale (PANSS)[10]. Subjects unable to speak Romanian, who were simultaneously enrolled in clinical trials or who had somatic comorbidities that may influence study outcomes were excluded.

### Sample Size

For this exploratory, transdiagnostic study we plan to recruit 300 participants distributed across five diagnostic groups (approximately 60 per group). This sample size is sufficient to detect small-to-medium between-group effects on continuous outcomes (e.g., negative symptom domain scores, reward and electrophysiological measures) using one-way or factorial ANOVA/linear models with an alpha of 0.05 and power of at least 0.80, while allowing adjustment for key covariates. It also provides adequate precision for multivariable regression and structural equation models examining the role of avolition and biological markers, maintaining a conservative ratio of at least 10–15 participants per estimated parameter. Given the primary focus on characterizing patterns and mechanisms rather than testing a single primary contrast, the chosen sample balances statistical power, model stability, and feasibility across the planned clinical, cognitive, biological, and electrophysiological analyses.

### Ethical Considerations

The study was approved by the Institutional Review Board Grigore T. Popa University of Medicine and Pharmacy Iasi Romania.

Participation in the study does not alter the patients’ ongoing clinical management. All clinical evaluations and psychometric assessments are performed as part of research procedures, and results are documented. Findings relevant to clinical care are communicated to the treating team and/or the participant’s legal guardian when appropriate.

### Data collection

All data are collected by trained study coordinators and clinical staff at the Socola Institute of Psychiatry, in compliance with institutional and ethical regulations for patient privacy and data security. Standardized clinical scales, neurophysiological assessments, and biological samples are obtained according to the study visit schedule to ensure consistency and reproducibility.

Negative symptoms are assessed using the Brief Negative Symptom Scale (BNSS) [11], administered by trained raters at multiple visits. To enhance reliability, interviews are audio– video recorded, capturing both patient responses and examiner behavior. These recordings are stored on secure, encrypted servers and are used for quality monitoring and inter-rater reliability checks. Additional validated instruments include the Positive and Negative Syndrome Scale (PANSS)[10], the Hamilton Depression Rating Scale (HAM-D)[12], the Udvalg for Kliniske Undersøgelser Side Effect Rating Scale (UKU)[13], the Barnes Rating Scale (BARS)[14], the Personal and Social Performance Scale (PSP)[15, 16], the Defeatist Performance Scale (DPAS)[17], the Temporal Experience of Pleasure Scale (TEPS) [18], and the Brief Assessment of Cognition in Schizophrenia (BACS)[19].

EEG recordings are conducted by a certified technician during hospitalization, following a standardized protocol. Each session includes an 8-minute resting-state acquisition, divided into alternating 2-minute blocks of eyes open and eyes closed. Data are processed for event-related potentials and spectral power analyses, aggregated at the group and symptom-profile levels. Raw EEG data are securely transferred to the CEMEX research infrastructure for storage and advanced processing.

Venous blood samples (3 mL, EDTA tubes) are collected at the final visit. Samples are kept at 4°C for a maximum of 4 hours prior to laboratory transfer. Following centrifugation, aliquots of 500 μL–1 mL are stored at –80°C. Genomic DNA is isolated and quality-controlled using Nanodrop, Qubit, and agarose gel electrophoresis. Genotyping will be performed via genome-wide microarray or whole genome sequencing, and polygenic risk scores (PRS) for persistent negative symptoms will be calculated. All biological samples are processed and stored at CEMEX.

### Data Security Protocols

All clinical, biological, and neurophysiological data are de-identified and stored on encrypted servers with restricted access. Consent forms and paper records are securely archived at CEMEX. Duplicate data entry and regular audits ensure integrity and completeness. Audio– video recordings of BNSS sessions provide an additional layer of quality control and facilitate inter-rater calibration.

All study staff undergo structured training and certification on the administration of scales and procedures prior to participant contact.

### Statistical Analysis

The statistical analysis plan focuses on characterizing negative symptoms across a transdiagnostic cohort with prominent negative symptoms. Analyses will begin with descriptive and multivariate methods to map the structure, severity, and distribution of negative symptom domains, followed by regression-based and structural equation approaches to examine the central role of avolition in shaping and maintaining other negative symptoms. Biological correlates of avolition will be explored through both targeted and data-driven analyses of peripheral biomarkers, neuroimaging indices, and electrophysiological measures. Additional models will investigate the pathophysiological mechanisms underlying distinct negative symptom domains, integrating behavioral, cognitive, and biological data. Reward processing—both anticipation and consummation—will be examined in relation to motivational deficits and their interaction with neurocognitive and social cognitive performance. Psychological networks of the interplay of symptoms will be examined as per network theory analysis methods [20, 21]. Electrophysiological profiles will be compared across symptom profiles, diagnostic groups, and treatment response categories, with predictive models assessing whether baseline neural signatures forecast clinical outcomes. Across all analyses, models will adjust for relevant covariates, apply appropriate corrections for multiple testing, and use robust methods to address missing data.

## Results

The initial version of the study protocol was developed in February 2024 and formally approved on May 8, 2024, with an updated version finalized on January 11, 2025. Participant recruitment and data collection began on July 1, 2024, at the Socola Institute of Psychiatry and are currently ongoing. Data collection is being conducted on a rolling basis during inpatient admissions. Interim data cleaning and quality control procedures are performed continuously throughout the recruitment period. Final data cleaning, statistical analyses, and interpretation of results will be conducted following completion of enrolment, with manuscript preparation planned thereafter.

## Discussion

Despite the substantial impact of negative symptoms on functional recovery and quality of life, their underlying mechanisms remain insufficiently characterized, particularly across diagnostic categories. This observational study addresses this gap by systematically examining the clinical, neurocognitive, electrophysiological, and biological correlates of negative symptoms, with a specific focus on avolition, across a transdiagnostic sample of individuals with severe mental illness and related conditions. By integrating standardized psychometric assessments with neurofunctional and polygenic risk measures, the study aims to improve the understanding of the pathophysiological foundations of negative symptoms. Findings from this research are expected to inform more precise diagnostic frameworks and support the development of targeted, mechanism-based therapeutic strategies.

## Anticipated Findings

Negative symptoms represent a major and persistent challenge across the spectrum of severe mental illnesses, contributing substantially to functional disability, poor quality of life, and limited response to current treatments. Despite their clinical relevance, negative symptoms remain insufficiently characterized from a transdiagnostic and biologically informed perspective. By systematically assessing negative symptoms across multiple diagnostic categories, the present study is expected to provide a more refined understanding of their phenomenology beyond traditional disorder-based frameworks.

Clarifying the role of avolition may therefore help explain the heterogeneity of negative symptom presentations across diagnostic groups, by integrating standardized psychometric assessments with electrophysiological measures and polygenic risk profiling, this study is expected to identify convergent biological and neurofunctional patterns associated with negative symptom severity.

The transdiagnostic nature of the sample represents a key strength of this investigation. Rather than focusing exclusively on schizophrenia, this study includes individuals with affective disorders and other conditions characterized by negative symptoms, thereby addressing an important gap in the literature.

Overall, the anticipated findings are expected to contribute to a more nuanced understanding of negative symptoms as biologically grounded, multidimensional phenomena.

## Limitations

This study has several limitations that should be acknowledged. First, participants were recruited from a single academic psychiatric center, which may limit the generalizability of the findings. Although the transdiagnostic nature of the sample enhances conceptual relevance, local clinical practices, patient characteristics, and referral patterns at the Socola Institute of Psychiatry may not fully represent other settings or populations. Multicenter studies will be required to confirm the robustness of the observed associations across different clinical contexts. Second, the assessment of negative symptoms relies partly on clinician-rated and self-report instruments. While validated scales and trained raters were used, self-report measures remain susceptible to recall bias, limited insight, and response variability, particularly in patients with cognitive impairment or severe psychopathology. In addition, symptom severity may fluctuate during hospitalization due to changes in clinical state or ongoing pharmacological treatment, potentially influencing assessment stability across visits. Third, despite standardized training procedures, the involvement of multiple raters introduces the possibility of inter-rater variability. Finally, participant attrition or exclusion during the assessment process may occur due to symptom instability, limited cooperation, or mistrust related to psychotic symptoms, which could introduce selection bias. These limitations should be considered when interpreting the findings. Nevertheless, the comprehensive multimodal approach and transdiagnostic focus of this study provide a strong foundation for future longitudinal and multicenter research aimed at refining the biological and clinical characterization of negative symptoms.

## Conclusions

This study will provide critical insights into the characterization and underlying mechanisms of negative symptoms across psychiatric disorders. By focusing on avolition, reward processing, and their interaction with neurocognitive and social cognitive deficits, it will help identify potential biological and electrophysiological markers of negative symptoms. The findings may guide the development of more precise assessment tools and inform novel therapeutic strategies, with broad translational impact for improving outcomes in individuals with serious mental illness and related conditions.

## Funding

This research was funded by Romania’s National Recovery and Resilience Plan (PNRR), Pylon III, Section I8. Development of a Program to Attract Highly Specialised Human Resources from Abroad in Research, Development and Innovation Activities, PNRR-III-C9-2023-I8, Project “ Modelling negative symptom domains neurobiology: a transdiagnostic, translational study”, code CF 46/ 28.07.2023.

## Conflicts of Interest

Authors Davidson and Rabinowitz are employed by Minerva Neuroscience.

## Data Availability

Not applicable.

## Notes

### Competing Interest Statement

Author Davidson serves as Medical Director and Rabinowitz as biostatistician for Minerva Neurosciences.

